# Fine-tuning large language models for effective nutrition support in residential aged care: a domain expertise approach

**DOI:** 10.1101/2024.07.21.24310775

**Authors:** Mohammad Alkhalaf, Jun Shen, Hui-Chen (Rita) Chang, Chao Deng, Ping Yu

## Abstract

**Purpose:** Malnutrition is a serious health concern, particularly among the older people living in residential aged care facilities. An automated and efficient method is required to identify the individuals afflicted with malnutrition in this setting. The recent advancements in transformer-based large language models (LLMs) equipped with sophisticated context-aware embeddings, such as RoBERTa, have significantly improved machine learning performance, particularly in predictive modelling. Enhancing the embeddings of these models on domain-specific corpora, such as clinical notes, is essential for elevating their performance in clinical tasks. Therefore, our study introduces a novel approach that trains a foundational RoBERTa model on nursing progress notes to develop a RAC domain-specific LLM. The model is further fine-tuned on nursing progress notes to enhance malnutrition identification and prediction in residential aged care setting.

**Methods:** We develop our domain-specific model by training the RoBERTa LLM on 500,000 nursing progress notes from residential aged care electronic health records (EHRs). The model’s embeddings were used for two downstream tasks: malnutrition note identification and malnutrition prediction. Its performance was compared against baseline RoBERTa and BioClinicalBERT. Furthermore, we truncated long sequence text to fit into RoBERTa’s 512-token sequence length limitation, enabling our model to handle sequences up to1536 tokens.

**Results:** Utilizing 5-fold cross-validation for both tasks, our RAC domain-specific LLM demonstrated significantly better performance over other models. In malnutrition note identification, it achieved a slightly higher F1-score of 0.966 compared to other LLMs. In prediction, it achieved significantly higher F1-score of 0.655. We enhanced our model’s predictive capability by integrating the risk factors extracted from each client’s notes, creating a combined data layer of structured risk factors and free-text notes. This integration improved the prediction performance, evidenced by an increased F1-score of 0.687.

**Conclusion:** Our findings suggest that further fine-tuning a large language model on a domain-specific clinical corpus can improve the foundational model’s performance in clinical tasks. This specialized adaptation significantly improves our domain-specific model’s performance in tasks such as malnutrition risk identification and malnutrition prediction, making it useful for identifying and predicting malnutrition among older people living in residential aged care or long-term care facilities.

## 1. Introduction

Malnutrition is a serious health problem with many negative health consequences for older people, such as a weakened immune system and impaired cognition [1]. It may also contribute to vulnerabilities of infections, anemia and other diseases [2–4]. Malnutrition has been identified as a key area for urgent review by the Australian government for residential aged care [5] with identification of poor nutrition and weight loss as important indicators measuring the quality of care in residential aged care facilities (RACF). Healthcare professionals are requested to regularly screen older adults for early detection of malnutrition [6, 7]. To date, the common malnutrition screening tools used at RACFs include Mini Nutritional Assessment (MNA) and Subjective Global Assessment (SGA). However, since using these tools are time-consuming, they were not consistently applied [7]. Predicting and addressing malnutrition can lead to better health outcomes and improved quality of life [8]. Thus, it is crucial to develop new methods to improve the efficiency and effectiveness in malnutrition detection. However, scarcity of reliable datasets detailing the nutritional intake, lack of domain-specific knowledge models, and the novelty of the transformer approach have been primary obstacles to the development of a malnutrition prediction model for older people.

### 1.1 Electronic health records

Electronic health records (EHRs) have been widely adopted in RACFs in Australia to document clients’ diagnosis, health assessment, nursing care plans, personal preferences, activities of daily living and care received [9]. The datasets in these EHRs can be classified as structured data and unstructured data. The structured data include client demographics and diagnosis that are recorded in structured tables. The unstructured data include nursing care plans, assessment records and free-text clinical notes [10]. Most information about clients in RACFs, including nutritional information, is recorded in unstructured progress notes in EHR. Since EHR data is captured real-time in the care service delivery process, models trained on EHR can be more readily applied to clinical practise [11]. This provides the opportunity for natural language processing (NLP) to extract insights from the unstructured data in EHR for aged care services.

### 1.2 Natural language processing

Recent advancements in artificial intelligence, more specifically NLP, have opened doors for extracting relevant information and automating clinical diagnoses and predictions using language models on patient EHR [12–14]. One of NLP’s recent advancements is the word embedding technique, which is a way of representing text as multi-dimensional vectors. Models such as GloVe [15] and word2vec [16] apply such text representation and have achieved promising results in different fields [17, 18]. However, these models lack context awareness, a core competency in text analysis.

### 1.3 Large language models

The emergence of the encoder-based large language models (LLM) such as Bert [19] and RoBERTa [20] have brought in positive disruption to the field of NLP. They utilize contextualized embeddings that account for both the prior and subsequent contexts of a token, adjusting its weight vector accordingly. By analyzing relationships between all pairs of words, LLMs introduce context-awareness, addressing the weakness of previous models. LLMs also have the ability to transfer previously acquired knowledge, thus are more efficient and have achieved state-of-the-art (SOTA) performance in many general downstream tasks with minimal to no need to modify architecture [21]. They can be further fine-tuned on specific corpus for specific-domain tasks. This has enabled them to be successfully fine-tuned for various complex applications. Previous studies have demonstrated that LLMs can be trained on medical corpus to achieve high reliability in medical diagnoses and predictions [22–24]. While LLMs have demonstrated their utility in extracting data from public health data sets, their practical application in specific clinical tasks within real clinical settings, using clinic data, remains limited [25, 26].

### 1.4 RoBERTa

RoBERTa is a robust encoder-based LLM that is further optimized from its predecessor BERT model for better performance on a variety of NLP tasks [20]. It has achieved SOTA performance after being trained with massive text data with increased parameters, larger batch size and learning rate. RoBERTa utilizes byte-level tokenization instead of word tokenization in BERT. In addition, it randomizes the masking place, which eliminates the chance for the model to memorize the training data. Previous studies found that encoder-based LLMs such as RoBERTa outperform or at least are as effective as decoder-based LLMs, e.g. ChatGPT, in classification task [27, 28]. RoBERTa’s architecture is highly suitable for fine-tuning on domain specific data sets. It has smaller model size, requires less computational power and memory, and often provides faster inference times compared to larger models like Llama model [29] or ChatGPT [30]. These make it more feasible for deployment in various health systems and devices [31]. Therefore, we choose RoBERTa as the candidate model for our task of generating knowledge about nutrition care in RACFs over other models.

### 1.5 Objective

Since, to date, there are no models that have been reported to be specifically fine-tuned for classifying and predicting malnutrition in older people, this study aimed to conduct NLP on free-text notes in the RAC EHR for two downstream tasks: (1) identifying malnutrition notes and (2) malnutrition prediction. We fine-tuned an encoder-based LLM, RoBERTa checkpoint, to produce a nutrition domain-specific LLM in Australian RAC setting. We evaluated the performance of our model in comparison with other baseline models, including BioClinicalBERT and RoBERTa. In addition, free text nursing notes within EHR often contain extensive and detailed documentation, however RoBERTa has a maximum sequence length limitation of only 512 tokens. To address this challenge, we developed a new method for processing long notes with length over the 512 token limit.

## 2. Methodology

### 2.1 Dataset

The dataset was obtained from 40 aged care facilities in the state New South Wales (NSW), Australia. Overall, 4,405 de-identified clients’ data was included in this analysis. The data was extracted from 1,616,820 notes of dietitians and nursing care staff recorded between Jan 2019 and October 2020, with an average number of 366 notes for each client. The Human research ethics approval for this study was granted by the Human Research Ethics Committee, the University of Wollongong and the Illawarra Shoalhaven Local Health District (Year 2020).

### 2.2 Data cleaning

All notes were cleansed of noise, including removing white spaces, special symbols and unwanted characters that do not contribute to the meaning of the text.

### 2.3 Overview of the methodology

Figure 1 depicts our NLP pipeline. It consists of three pathways: Path 1, fine-tuning a domain specific LLM; Path 2, finetuning a malnutrition note identification model; Path 3, finetuning malnutrition prediction model.

**Fig. 1:**
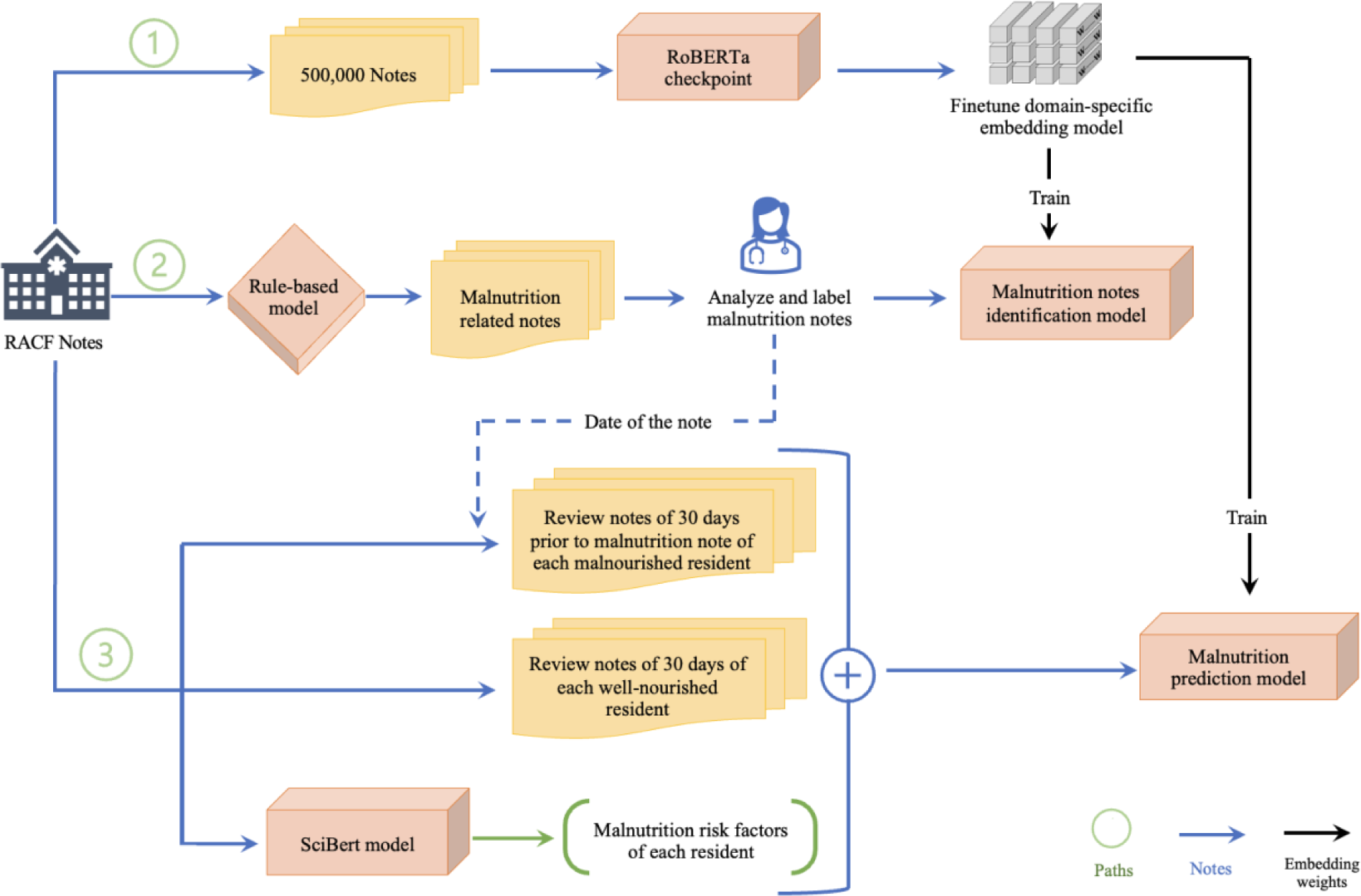
An overview of the model development pathway. Path 1: Fine-tuning a RAC domain-specific model Path 2: Fine-tuning a malnutrition note identification model Path 3: Fine-tuning a malnutrition prediction model

### 2.4 Path 1: Fine-tuning domain-specific embedding model

#### 2.4.1 Dataset construction

We randomly selected 500,000 free-text nursing notes with an average token length of 64. The training dataset included 21,969,925 words, which we considered adequate for domain-specific fine-tuning as more notes do not necessarily lead to better results [32]. We then extracted the raw text to a single text file and processed it into chunks of 512 tokens. This chunking procedure resulted in a training set containing 62,273 text chunks (rows).

#### 2.4.2 Model fine-tuning

The weights of the baseline RoBERTa model checkpoint (“roberta-base”) downloaded from the Huggingface transformer library [33] contain substantial information regarding the English language corpus. This substantially reduces the fine-tuning time adapting RoBERTa to our specific task than training a model entirely from scratch. However, the corpus of nursing progress notes contains many RAC domain-specific terms, abbreviations and unconventional expressions that do not present in general English, which could affect the performance of the baseline RoBERTa model. Therefore, we chose to train a RAC domain-specific model initialized from RoBERTa on our nursing note dataset to improve the model’s ability to understand the words and phrases used in the RAC nursing corpus. The task for the model is to predict words randomly masked out of an input chunk. The knowledge of the resulting model can be transferred and further trained with an additional output layer to create models for various downstream tasks.

Tokenization was conducted on the nursing text corpus using a pre-trained byte-level tokenizer to fit with the RoBERTa model. We randomly split the dataset into 80% training and 20% validation sets. Then, we set the masking probability to 15% of the words in each input sequence, like the original RoBERTa training. We used whole word masking instead of token masking for better results [34] (Supplementary Table S1). In addition, we randomize the masking with each batch to avoid over-memorization. After that, we trained the model with the following hyperparameters: learning rate of 1e-4, batch size of 32, and weight decay of 0.01. The model was trained until validation loss started to converge (Supplementary Figure S1). The embeddings of this model were then utilized for the two downstream tasks: malnutrition note identification and malnutrition prediction.

### 2.5 Path 2: Downstream task 1: Fine-tuning a malnutrition note identification model

#### 2.5.1 Dataset construction

Transformer models need a considerable amount of labelled data to boost their performance. However, to the best of our knowledge, there is not any publicly available, malnutrition-labelled data; therefore, we engaged three nursing domain experts to build a malnutrition-specific labelled dataset. To accomplish this, we developed process to identify and label records with malnutrition [35]. We first constructed a rule-based model to identify all malnutrition notes in the dataset. Using these rules, we extracted 2,474 notes belonging to 1,283 clients. Manual analysis and screening of all extracted notes identified 196 notes that did not fit the malnutrition definition, but either reported planned weight loss or invalid weight recording due to scale or typing errors. At the end, our manually labelled ground truth training dataset contained 2,278 notes reporting malnutrition (labelled: 1) and 15,000 notes with normal nutrition status (labelled: 0).

#### 2.5.2 Model finetuning

We further fine-tuned our model with the embeddings from the RAC domain-specific LLM that we built in Path 1 to identify notes related to malnutrition (see Figure 1). We divided the dataset into 85% training and validation datasets, and 15% for hold-out testing set. The hyperparameters included learning rate of 3e-5, batch size of 16, weight decay of 0.01 and 50% of dropout rate. We used binary cross-entropy loss with positive weights and the mean pooling output of the last hidden state.

### 2.6 Path 3: Downstream task 2: Fine-tuning a malnutrition prediction model

#### 2.6.1 Dataset construction

The dataset for this task consisted of the original weekly nursing review notes and the malnutrition risk factors extracted from these notes. Since malnutrition is a health condition that develops over time, to capture each client’s health changes over time, we approached this task as a time series data analysis by extracting weekly review notes of each malnourished client recorded in the 30 days prior to the onset of malnutrition. We organized each client’s notes chronologically, with the earliest note appearing first in the sequence and the most recent note appearing last. We followed the same procedures to organize data for clients without malnutrition.

In addition to text-based notes, we also extracted malnutrition risk factors for each client from the notes using the SciBert model for named entity recognition with UMLS linker [36]. In our previous study, we identified 46 malnutrition risk factors in each client’s notes such as poor appetite, suboptimal oral intake and dysphagia [35]. Moreover, we applied a negation technique to distinguish whether a factor mentioned in a note was confirmed or negated [37]. For instance, if a note has the following sentence “no sign of cancer”, the algorithm will correctly identify this as a negation and not a confirmed factor. Finally, we combined each client’s notes and the risk factors into one file. We added the notes as raw text data and the risk factors as one-hot encoding tensor, with ‘0’ indicating the absence of the factor and ‘1’ indicating its presence. After that, we used notes and factors as the dataset for the malnutrition prediction model.

#### 2.6.2 Model finetuning

Our model was initialized from the RAC domain-specific model fine-tuned in Path 1. The training dataset consisted of 862 aggregated notes (rows) of malnourished clients and 2,298 aggregated notes (rows) of well-nourished clients. We split the data into 85% for training and validation, and 15% for hold-out testing. Hyperparameters included a learning rate of 3e-5, batch size of 16, weight decay of 0.01 and 50% dropout rate. We used binary cross-entropy loss with positive weights. We concatenated the output with the structured data (malnutrition risk factors). We added to the concatenated data a fully connected layer, with a Sigmoid activation function applied to obtain the final output (Supplementary Figure S2).

#### 2.6.3 Addressing the 512 maximum length challenge

In this downstream task, as opposed to Task 1, the notes were longer due to the inclusion of information gathered over a four-week period. Therefore, we encountered long notes spanning a four-week duration, with an average 644 token length (95% confidence interval:627.17 - 663.31). Therefore, the length of certain records exceeded the maximum sequence length accepted by RoBERTa and BERT, which is 512 tokens.

For these long records, we truncated and padded the text sequence into equal 512 token parts. Each part starts with a start sequence token and concludes with an end sequence token. Padding token was added if the last part has less than 512 tokens. Attention masks were also manually added as (1), informing the model to pay attention to the token, or (0), suggesting the model to ignore the token.

In the model forward function, the last hidden state embeddings of each token, generated by the model, are selectively emphasized through an attention mask. Then the sum of the masked embeddings is calculated. Only tokens with an attention mask value of 1 are considered; tokens with an attention mask of 0, which indicated padding token, are ignored. In addition, the model keeps track of the number of tokens. To capture the main ideas of a note, for short notes, embeddings are averaged across all tokens (Figure 2A). Conversely, as longer notes are divided into several parts with equal length of 512 tokens, embedding tokens are aggregated across all parts (Figure 2B).

**Fig. 2:**
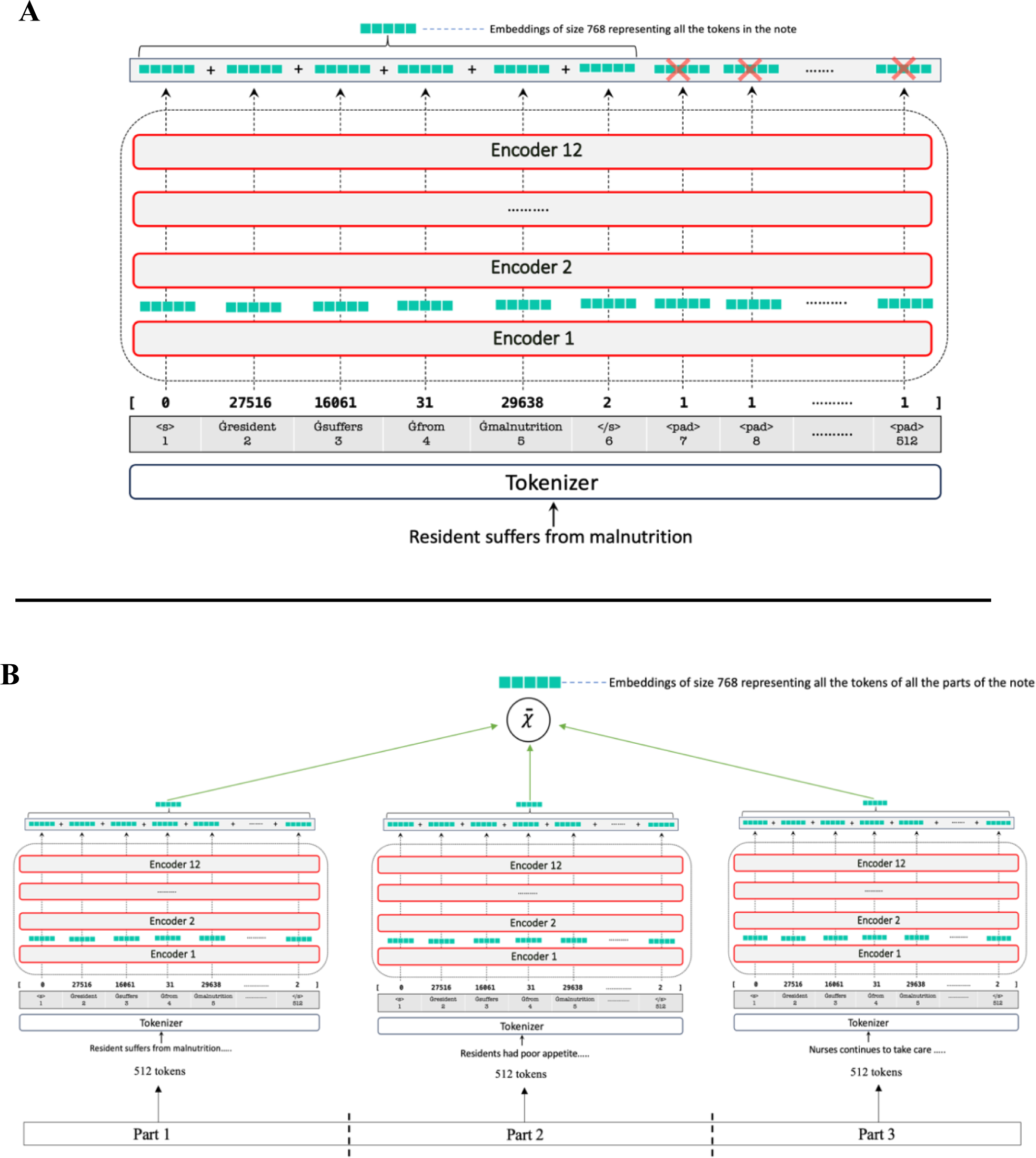
Methods developed for processing long notes with more than 512 tokens. (**A**) Example of a nursing note with sequence length less than 512 tokens; (**B**) Example of a nursing note with sequence length of 1536 tokens. This note is truncated into 3 parts each with a sequence of 512 tokens.

All tasks were evaluated using precision, recall, F1-score, specificity, the area under the precision-recall curve (AUPRC) and the area under the receiver operating characteristic curve (AUROC). To better assess the model’s robustness, generalizability and avoid finetuning instability [38], we performed 5-fold cross-validation in each downstream task. We kept the number of epochs in each fold to a low number (four) to avoid the possibility of overfitting the data. In each fold, the model with the least validation loss was utilized for testing on the test dataset. We calculated the cross-validation performance by taking the average of the k performance estimates of all measures (F1-score, recall, etc.) obtained from the testing sets using the arithmetic mean. Additionally, we calculated the confidence interval for each measure.

This study was implemented using Python 3.10.11, PyTorch 2.0.1, transformers 4.29.2 (Huggingface) and Scikit-learn 1.2.2. Our models were trained on the NVIDIA Tesla T4-16GB graphics processing unit (GPU).

**Table 1:** The proportion of malnourished clients in the studied population (n = 4,405)

|  | Well-nourished (n=3,204) | Malnourished (n = 1,201) |
| --- | --- | --- |
|  | Mean (SD) | Mean (SD) |
| Age | 85.2 (8.9) | 85.1 (8.9) |
| Female | 2,071 (74%) | 726 (26 %) |
| Male | 1,133 (70%) | 475 (30 %) |

## 3. Result

We compared the results of our domain specific LLM to the baseline models, RoBERTa, and BioClinicalBERT. In the first downstream task (malnutrition note identification), LLMs had very similar results, with RAC domain specific LLM producing a slightly better F1-score of 0.966, followed by RoBERTa, which achieved a comparable F1-score of 0.964. Then, BioClinicalBERT had an F1-score of 0.960 (Table 2).

**Table 2:**
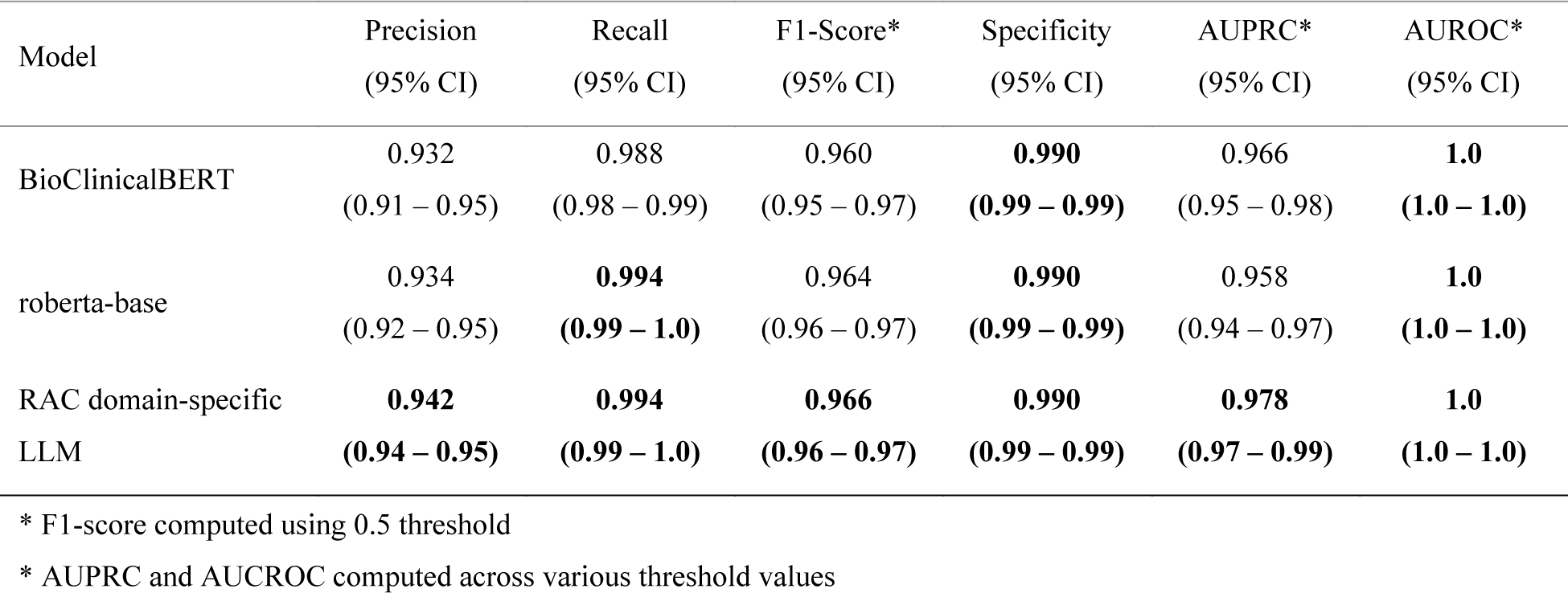
Performance of the five machine learning models on the task of identifying malnutrition notes.

In the second downstream task of malnutrition prediction, LLMs demonstrated superior performance to the older techniques. RAC domain specific LLM with the risk factor layer was the top-performing model with an F1-score of 0.687. It was followed by our domain specific LLM without the risk factors, which had an F1-score of 0.655. Next, RoBERTa had an F1-score of 0.614. Then, BioClinicalBERT achieved an F1-score of 0.582 (Table 3). Supplementary Figures (S3 – S9) show the area under the curve plots for each model.

**Table 3:** Results of the malnutrition prediction model.

| Model | Precision<br>(95% CI) | Recall<br>(95% CI) | F1-Score*<br>(95% CI) | Specificity<br>(95% CI) | AUPRC*<br>(95% CI) | AUROC*<br>(95% CI) |
| --- | --- | --- | --- | --- | --- | --- |
| BioClinicalBERT | 0.554<br>(0.51 – 0.59) | 0.617<br>(0.54 – 0.70) | 0.582<br>(0.54 – 0.62) | 0.787<br>(0.74 – 0.84) | 0.613<br>(0.56 – 0.67) | 0.771<br>(0.74 – 0.80) |
| roberta-base | 0.579<br>(0.50 – 0.66) | 0.662<br>(0.59 – 0.74) | 0.614<br>(0.58 – 0.65) | **0.789**<br>**(0.71 – 0.87)** | 0.677<br>(0.66 – 0.70) | 0.803<br>(0.79 – 0.82) |
| RAC domain-specific<br>LLM | 0.592<br>(0.50 – 0.68) | 0.751<br>(0.64 – 0.86) | 0.655<br>(0.62 – 0.69) | 0.766<br>(0.63 – 0.90) | 0.734<br>(0.67 – 0.80) | 0.843<br>(0.82 – 0.87) |
| RAC domain-specific<br>LLM + risk factors | **0.615**<br>**(0.53 – 0.70)** | **0.790**<br>**(0.71 – 0.87)** | **0.687**<br>**(0.65 – 0.72)** | 0.780<br>(0.67 – 0.89) | **0.735**<br>**(0.68 – 0.79)** | **0.858**<br>**(0.83 – 0.88)** |

\* F1-score computed using 0.5 threshold
\* AUPRC and AUCROC computed across various threshold values

## 4. Discussion

The aim of this study was to develop an encoder-based RAC domain specific LLM to accurately identify clients with malnutrition in EHR and develop a model capable of predicting malnutrition in older people one month before its onset. We first utilized our RAC domain-specific LLM, initialized from the well-known RoBERTa model and further fine-tuned on nursing progress notes. Afterwards, we employed the embedding weights generated from the proposed model for two subsequent downstream tasks. We compared the performance of different parameters on three models, BioClinicalBERT, roberta-base and our domain-specific LLM. The results demonstrated the advantage of utilizing domain-specific embeddings. It is worthy to note that this is the first study to utilize LLMs on free text nursing notes to predict malnutrition in older people, although it has been a health risk that has long plagued the care staff members and has casted a negative impact on care quality [1]. This study has also developed a method for processing long notes (> 512 tokens), which is crucial and particularly relevant in health contexts where it is typical to encounter long and detailed documentation.

For the first downstream task, there have been very few attempts to identify malnutrition notes in EHR in the literature. One attempt to classify malnutrition notes applies conditional random fields technique to nursing notes [39]. However, the study reported that the model performed poorly on malnutrition and had a low F1-score of 0.39. The authors of the study stated that classifying malnutrition notes was very challenging which led to the low accuracy of their model. Another attempt is our previous work to classify the malnutrition notes using a rule-based model which achieved a high-level performance; however, the development of the rule-base method was time-consuming and labor-intensive [35]. To address this limitation, we adopted a specific domain LLM in this study. The process of fine-tuning and evaluating the model was much more efficient than the rule-based method.

In accordance with the previous reports [22, 40, 41], LLMs significantly outperformed other comparative models with our RAC domain-specific LLM achieving an F1-score of 0.966. However, all LLMs models yielded comparable performance in this task. We argue that this can be attributed to the notable difference in notes between the positively labelled and negatively labelled instances in the dataset compared to those in the second task.

For the second downstream task of malnutrition prediction, to our knowledge, this is the first study in predicting malnutrition for older people in RACFs applying a transfer learning approach to EHR. Once again, our RAC domain-specific LLM notably outperformed other LLMs and had the highest F1-score of 0.655. This illustrates the importance of fine-tuning a foundational LLM on a domain-specific corpus. In addition, combining a layer of structured data with the output of the note-based model increased the performance of the model, as evidence by an increased F1-score (0.687). Despite that, the model did not achieve a high F1-score like in the first task, which is arguably because malnutrition is a health risk that is influenced by various factors, many of which are prevalent in all clients and are not specific for clients with malnutrition. Therefore, accurately predicting malnutrition is still a complex and challenging mission [6, 42].

Our findings in this study will be practically and clinically important for the malnutrition management of older people in RACFs. We would also stress that our LLM is not a replacement for already existing reliable screening tools. Instead, it could be incorporated into the nursing care process to help nurses and clinicians efficiently identify clients at risk of malnutrition by automatic screening of their EHR data. This will enable them to implement the tailored malnutrition prevention and intervention actions accordingly. In addition, our study demonstrates the feasibility of using a robust, well-known LLM such as RoBERTa can facilitate researchers to produce the optimal model for various downstream tasks.

The area of LLMs is evolving at a rapid pace; recently, the decoding models such as GPT-3.5 and GPT-4, though with widely doubted hypes, have revolutionized the whole field of NLP. However, there remains a paucity of usable models for health care systems [26]. We intend to compare the prediction ability of our encoder-based LLM with one of the more recent and advanced decoder-based LLMs such as Llama 2 [29]. We will also further gather nursing notes to improve the performance of the model, particularly for the second task.

### 4.1 Limitation

This study has two notable limitations. Firstly, LLMs typically require substantial training data to achieve high levels of accuracy [15]. In the case of our malnutrition prediction task, the dataset size is relatively modest, potentially impacting the model’s predictive performance. The availability of a larger dataset could lead to improved accuracy and more robust predictions. Secondly, although our models are trained on data sourced from a diverse array of 40 RACFs, it’s crucial to acknowledge that these facilities are all part of a single organization. Consequently, the applicability of our models might be constrained when used in RACFs with differing strategies and guidelines for electronic data collection. The lack of diversity in institutional practices could potentially hinder the models’ generalizability to a broader range of settings.

## 5. Conclusion

To address the critical malnutrition issue in older people, we proposed an encoder-based RAC domain-specific LLM that is fine-tuned from the foundation LLM, RoBERTa model, on RAC domain-specific nursing text. The resulted embeddings were successfully utilized for two downstream tasks: malnutrition note identification and malnutrition prediction. Our findings demonstrate that fine-tuning a foundation LLM with domain-specific corpus can improve the performance of the foundation models. In addition, combining risk factors as a structured data with a text model enhance model performance. This study also developed a method to truncate long text into parts that fit into the 512-token limit of RoBERTa model.

## Data Availability

All data used in the present study are sensitive and cannot be shared

## Author contribution

Design of work: Mohammad Alkhalaf and Ping Yu. Data analysis and interpretation: Mohammad Alkhalaf and Ping Yu. Drafting of the manuscript: Mohammad Alkhalaf and Ping Yu. Critical review and revision: all authors.

## Acknowledgments

The first author, Mohammad Alkhalaf, is supported by a full PhD scholarship from Qassim University, Saudi Arabia. The authors are grateful for the aged care organization that shared the de-identified electronic health records, which provided the opportunity to conduct this significant research project.

## Statements and Declarations

The authors declare that they have no conflicts of interests that could have appeared to influence the work reported in this paper.

## Statement on funding

No funding was received for conducting this study.

## Ethical approval

The Human research ethics approval for this study was granted by the Human Research Ethics Committee, the University of Wollongong and the Illawarra Shoalhaven Local Health District (Year 2020).

## 6. Supplementary materials

**Supplementary Table S1:**
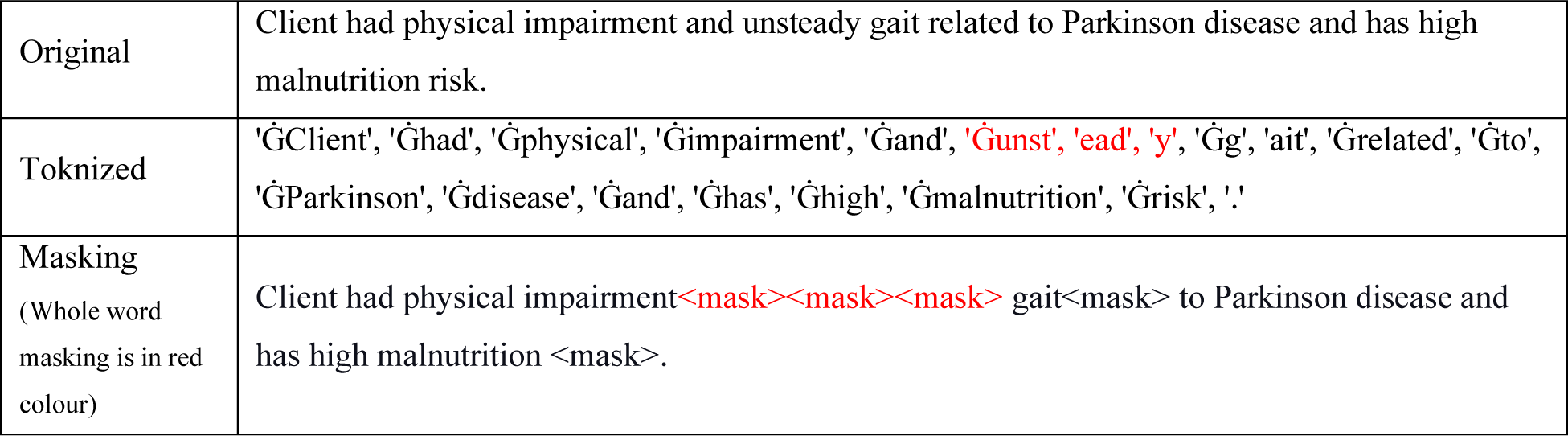
Example of tokenization and 15% whole word masking.

**Supplementary Figure S1.**
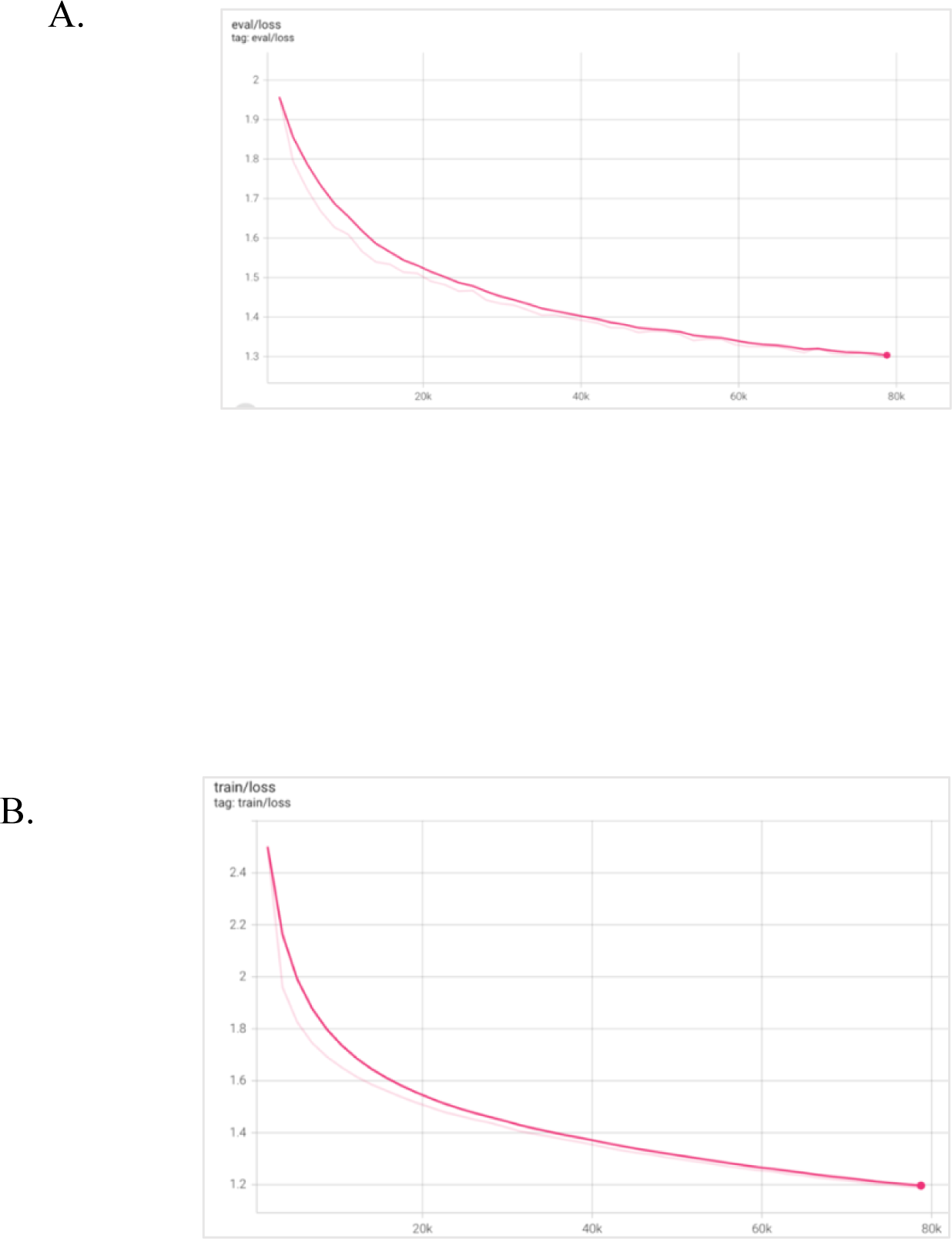
The RAC domain-specific LLM training, (A) validation loss, (B) train loss

**Supplementary Figure S2.**
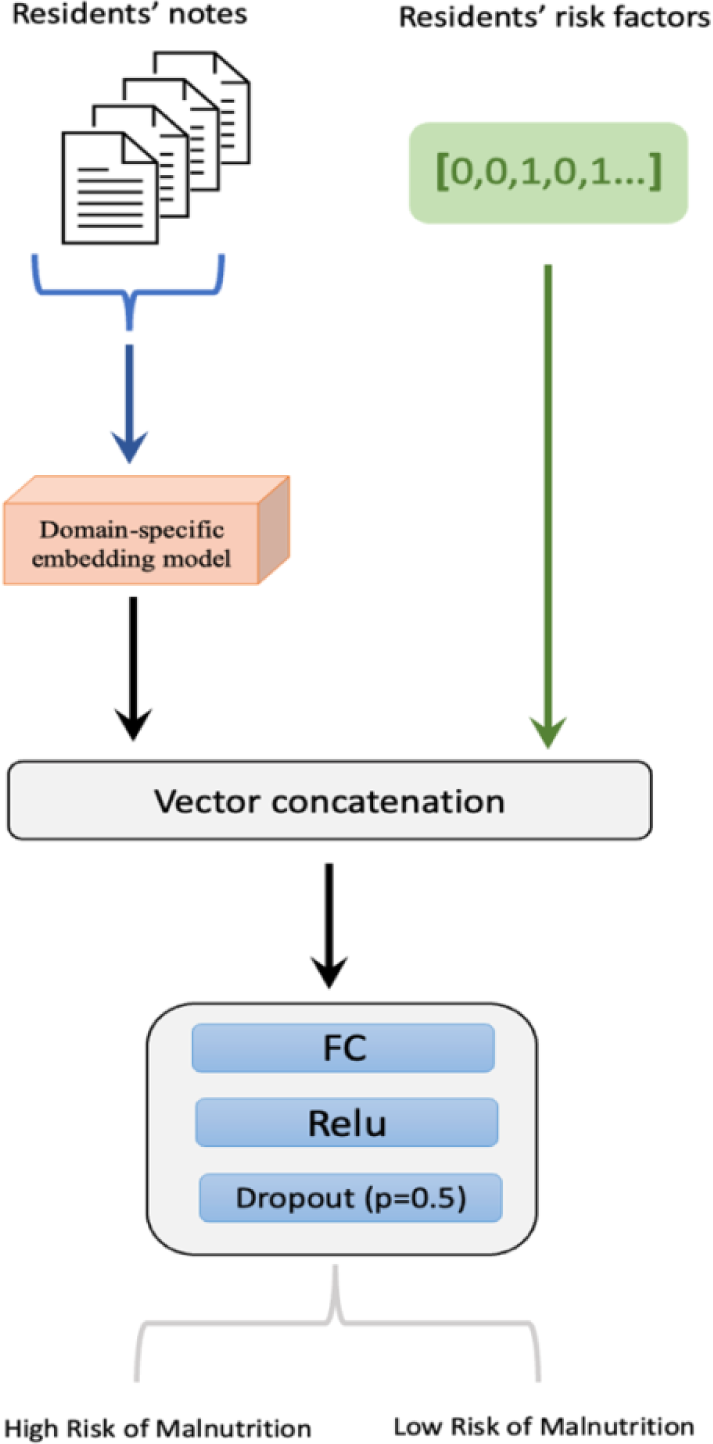
The Malnutrition prediction model

**Supplementary Figure S3.**
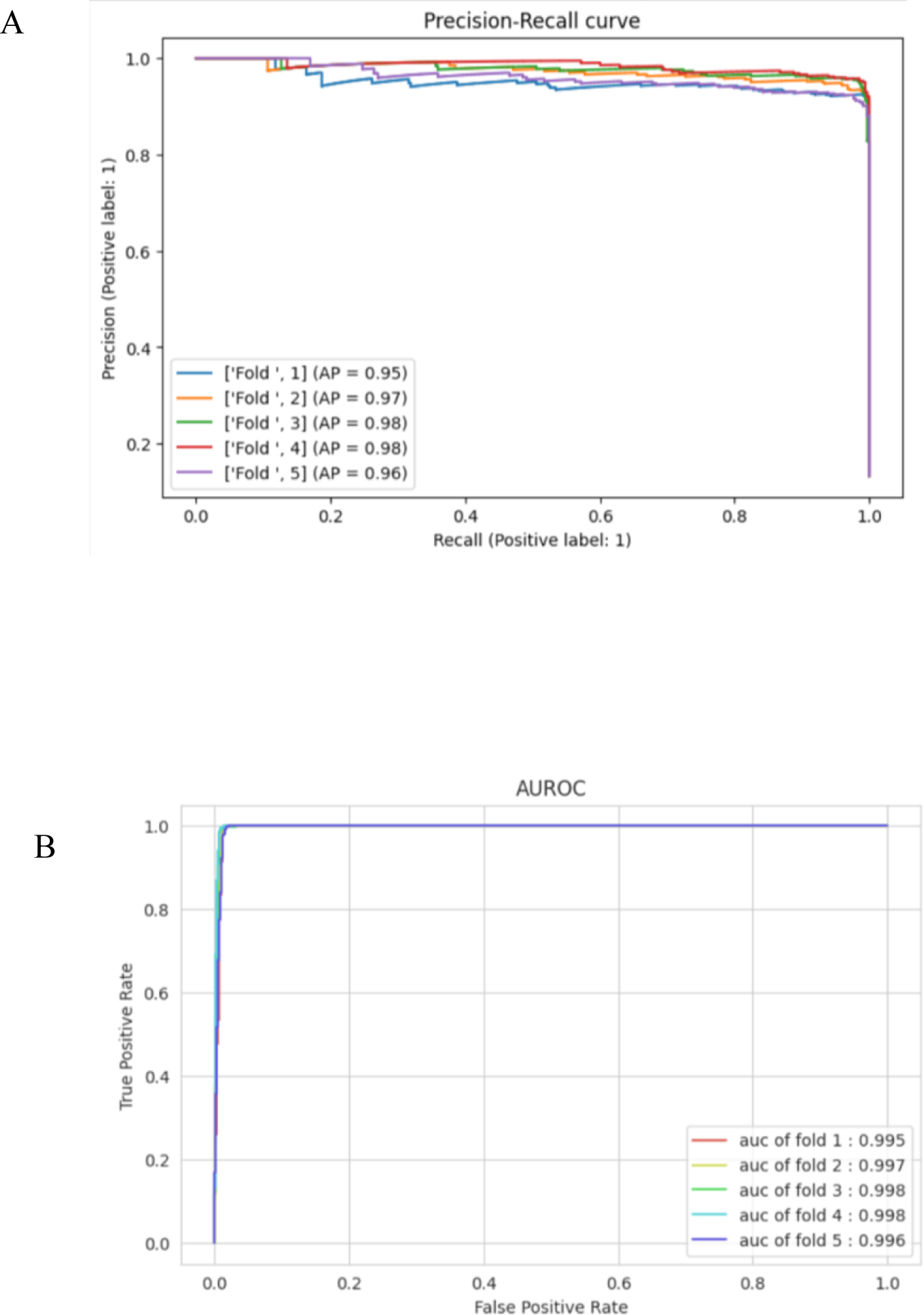
AUPRC (A) and AUROC (B) of malnutrition note identification model - BioClinicalBERT

**Supplementary Figure S4.**
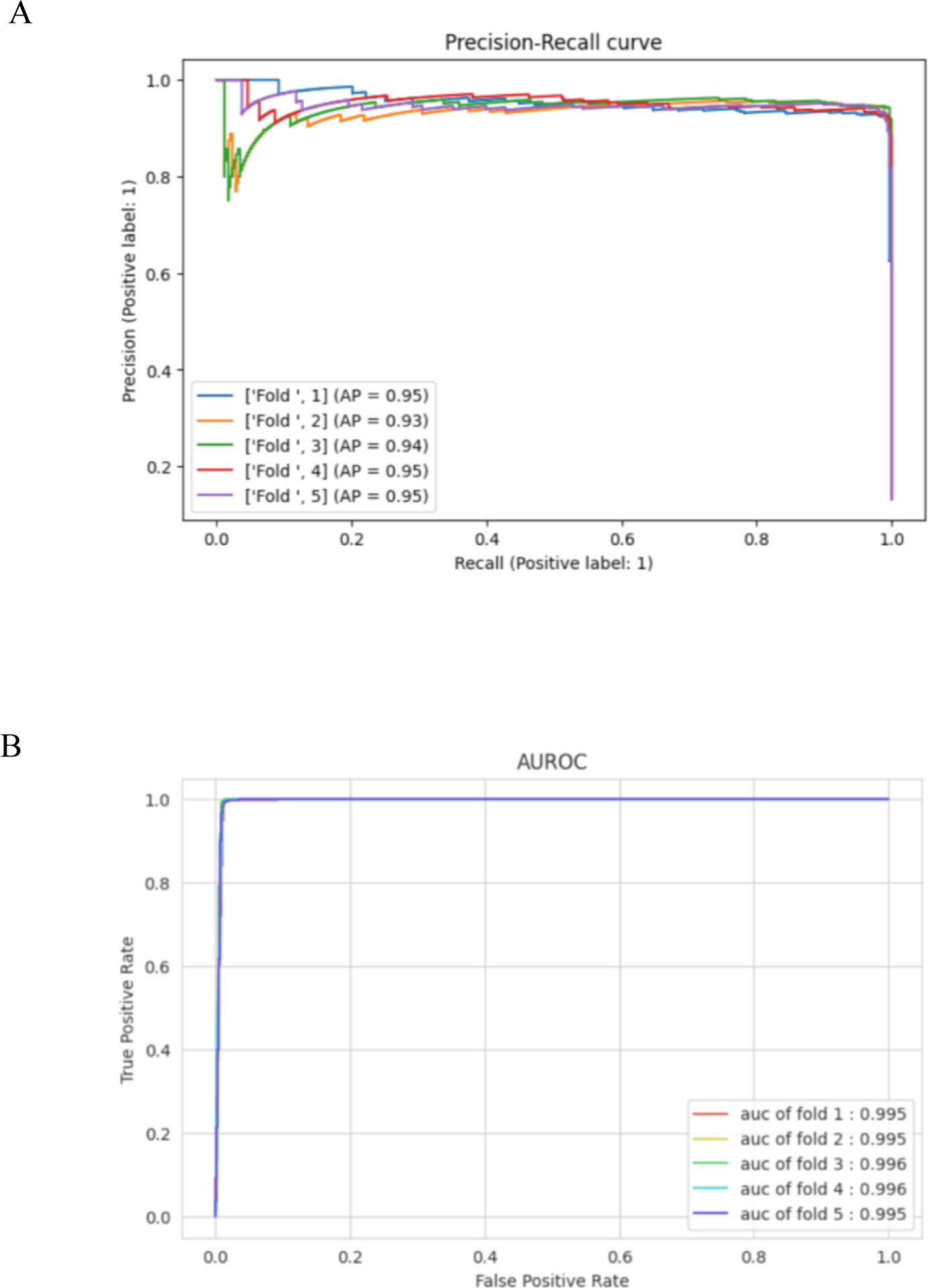
AUPRC (A) and AUROC (B) of malnutrition note identification model – RoBERTa base

**Supplementary Figure S5.**
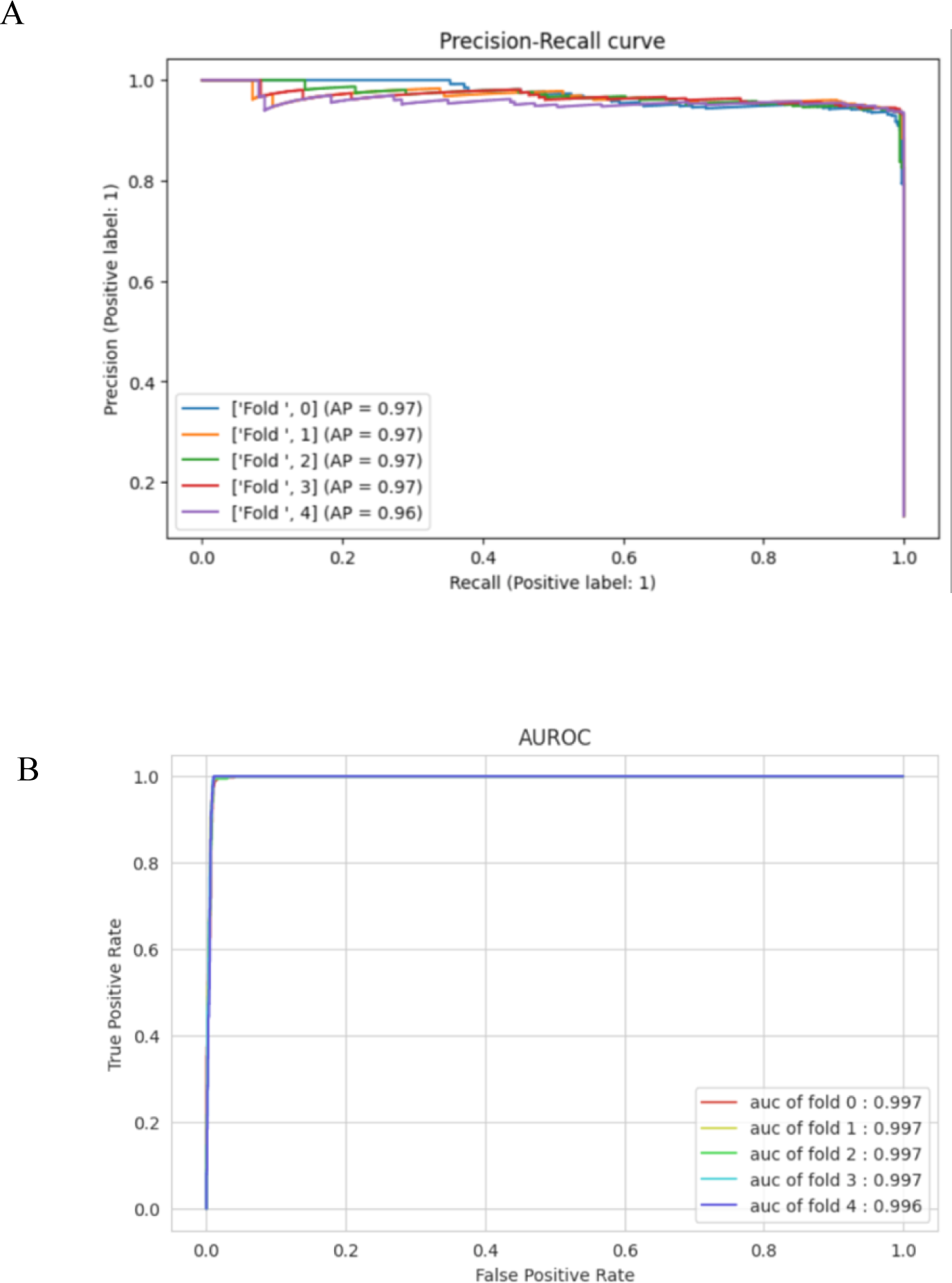
AUPRC (A) and AUROC (B) of malnutrition note identification model - RAC domain-specific LLM

**Supplementary Figure S6.**
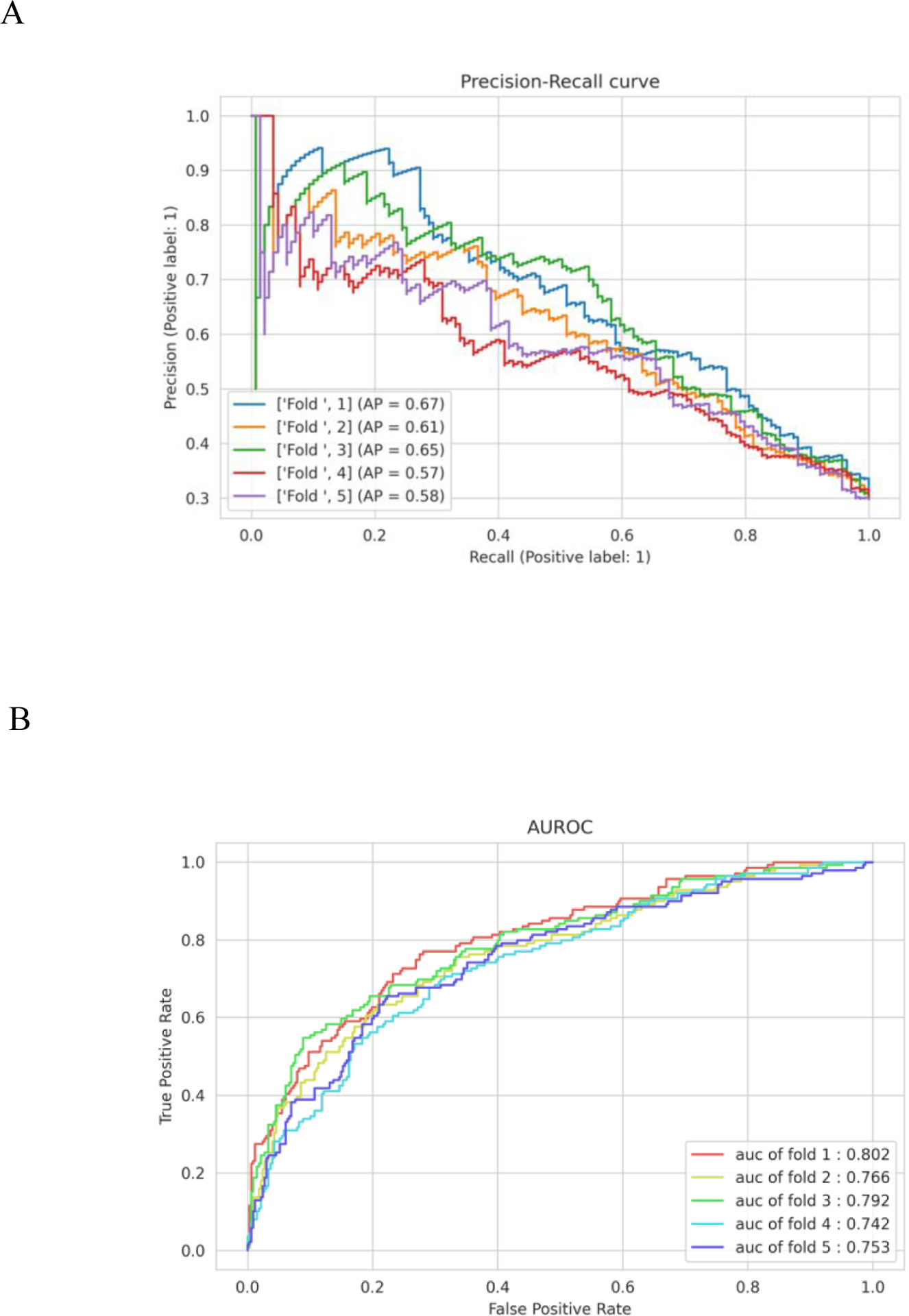
AUPRC (A) and AUROC (B) for malnutrition prediction model - BioClinicalBERT

**Supplementary Figure S7.**
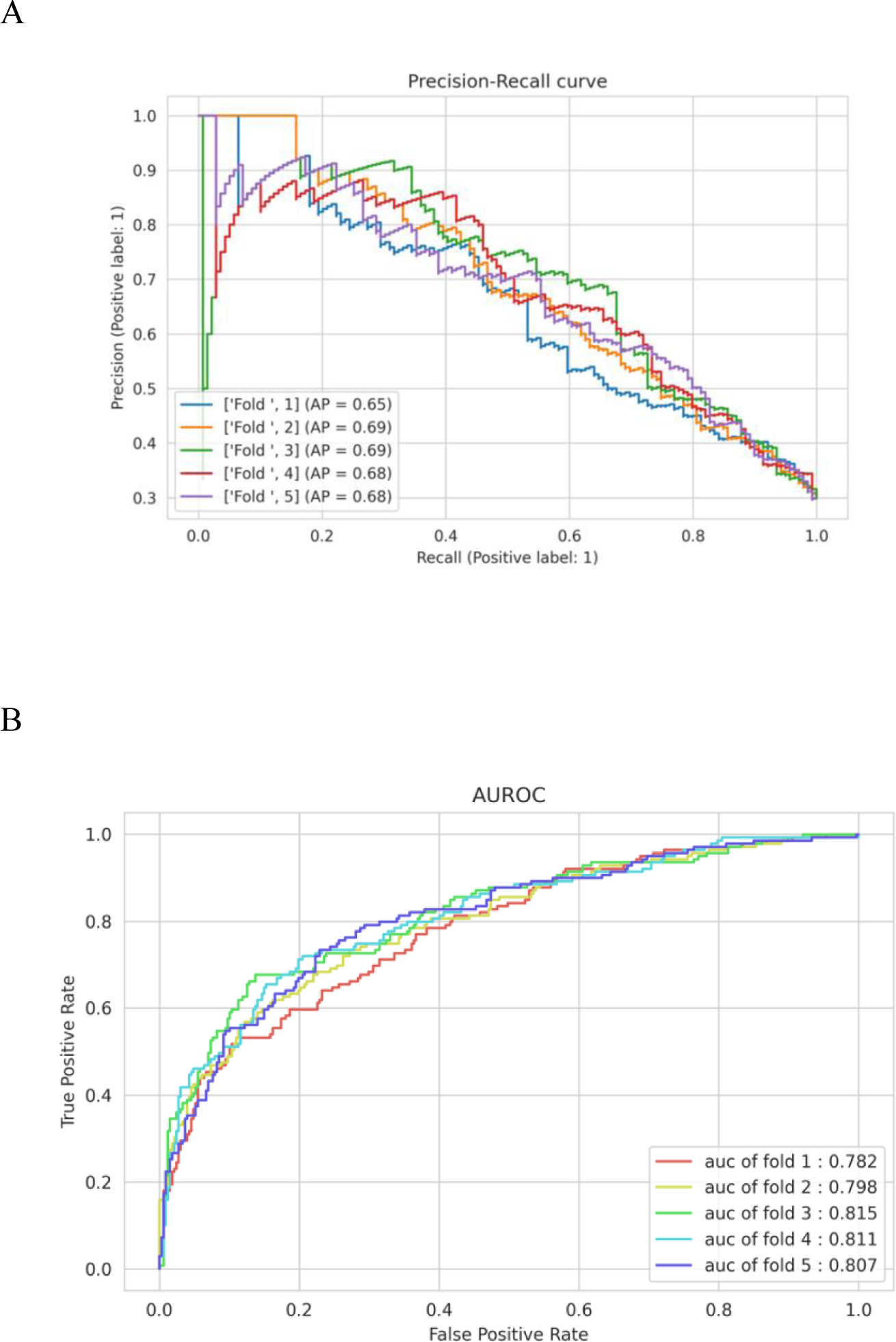
AUPRC (A) and AUROC (B) for malnutrition prediction model - RoBERTa base

**Supplementary Figure S8.**
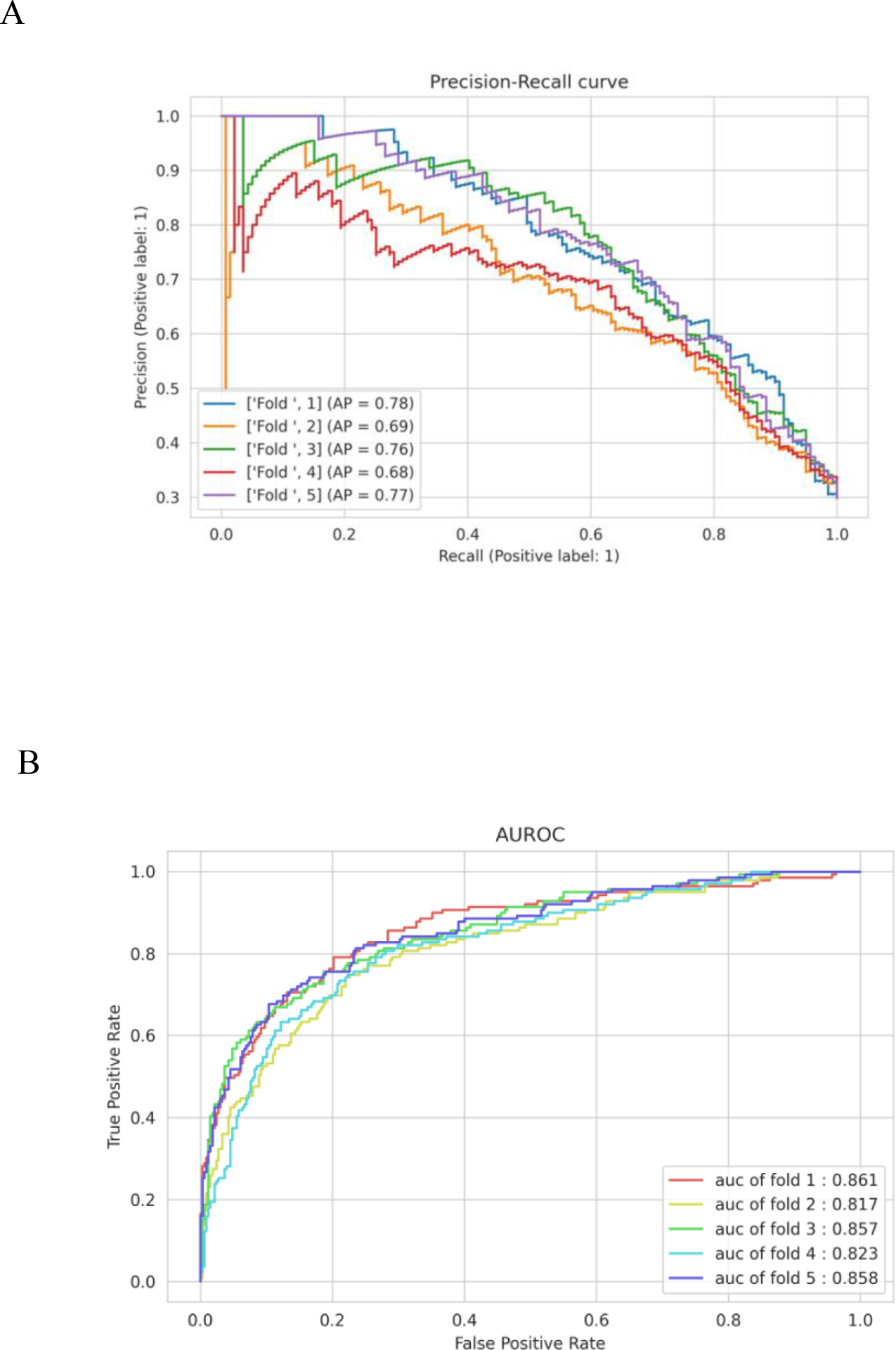
AUPRC (A) and AUROC (B) for malnutrition prediction model - RAC domain-specific LLM

**Supplementary Figure S9.**
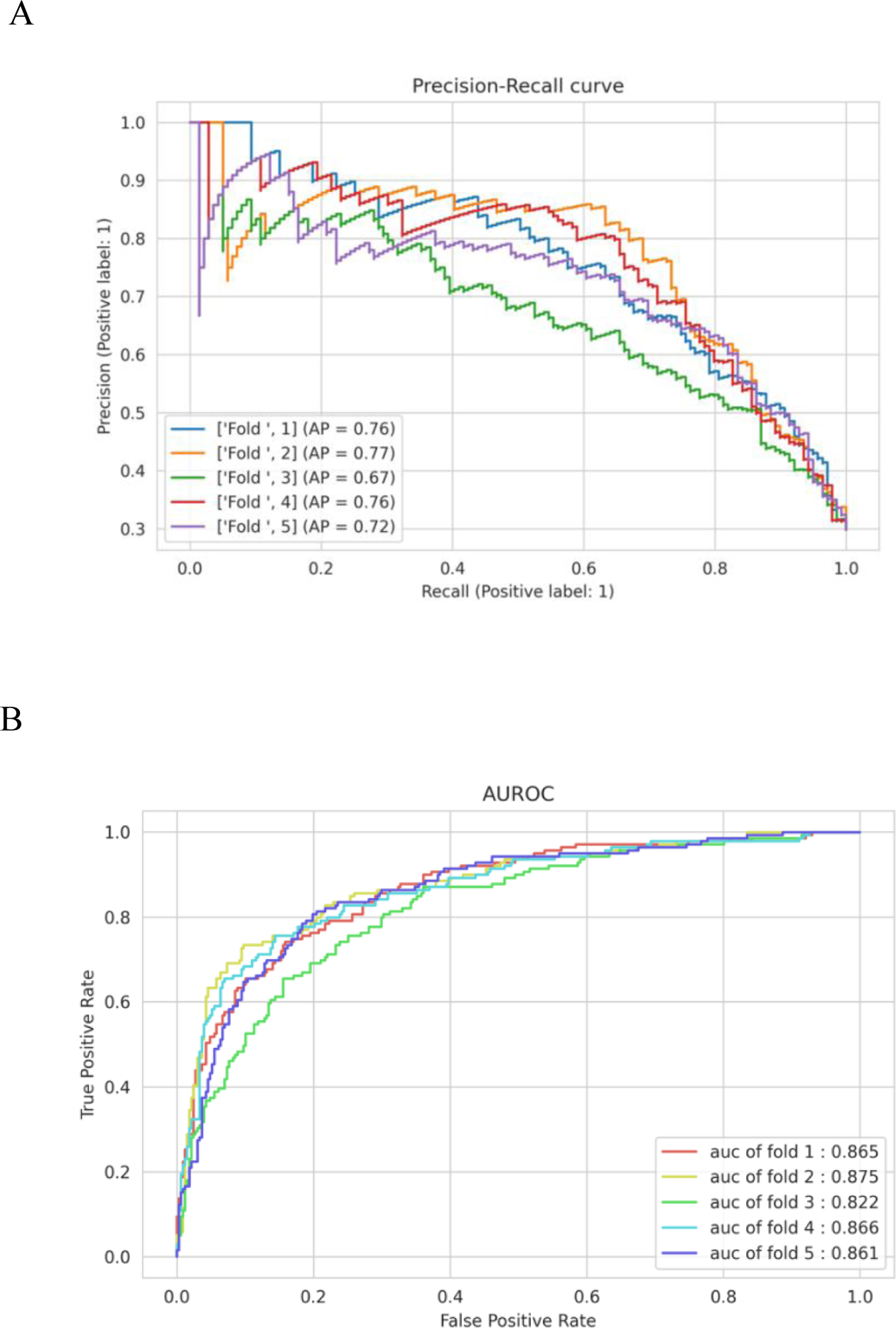
AUPRC (A) and AUROC (B) for malnutrition prediction model - RAC domain-specific LLM with risk factor layer

## Notes

### Competing Interest Statement

The authors have declared no competing interest.

### Funding Statement

This study did not receive any funding

